# Characteristics associated with COVID-19 vaccination status among staff and faculty of a large, diverse University in Los Angeles

**DOI:** 10.1101/2021.09.29.21264315

**Authors:** Michele Nicolo, Eric Kawaguchi, Angie Ghanem-Uzqueda, Andre E. Kim, Daniel Soto, Sohini Deva, Kush Shanker, Christopher Rogers, Ryan Lee, Yolee Casagrande, Frank Gilliland, Sarah Van Orman, Jeffery Klausner, Andrea Kovacs, David Conti, Howard Hu, Jennifer B. Unger

**Affiliations:** Department of Population and Public Health Sciences, Keck School of Medicine, University of Southern California, Los Angeles California; Family Medicine, Keck Medicine of USC, Los Angeles, California; Keck School Medicine of USC, University of Southern California, Los Angeles California; Keck Medical Center of USC, Los Angeles, California

## Abstract

**Objective:** This study examined characteristics associated with being unvaccinated among a sample of university staff and faculty prior to university campus reopening for in-person learning in spring-summer 2021.

**Methods:** Staff and faculty responded to an email invitation to complete an online survey. Survey questions included demographic data (race/ethnicity, age, sex), COVID-19 knowledge and behaviors, employment specific data including division and subdivision (healthcare vs. non-healthcare related division); and self-reported vaccination status. A multivariable logistic regression analysis was performed to determine significant characteristics associated with the likelihood of being unvaccinated for COVID-19.

**Results:** Participants identifying as Asian and Asian American, Hispanic/Latinx or Multicultural/Other had greater odds of being unvaccinated compared to Non-Hispanic White participants. Other characteristics associated with greater likelihood of being unvaccinated included working as university staff member (vs. faculty), older age, decrease in income, inability to work remotely and not traveling outside of Los Angeles area. Political affiliation as an Independent or as something else were more likely to be unvaccinated compared to participants identifying as Democrat.

**Conclusions:** Findings suggest several factors associated with racial and social disparities may delay the uptake of COVID-19 vaccination. This study highlights the need for targeted educational interventions to promote vaccination among university staff and faculty.

## Background

COVID-19 vaccination is the best method for reducing morbidity and mortality.[1] Despite variants of COVID-19 becoming more transmissible, ∼30-50% of vaccine eligible individuals in the U.S. remain unvaccinated.[2-6]. Health disparities, mistrust of government and medical institutions, misinformation, political beliefs, and trepidation over unknown long-term side-effects are often cited reasons for vaccine hesitancy.[7-14]

As higher education institutions resume in-person learning and on campus activities, the likelihood of COVID-19 outbreaks will continue to be a concern for college and university communities.[15] Successful campaigns encouraging vaccination will depend on understanding vaccine hesitancy. To investigate reasons for lower than optimal vaccination rates among university employees, the current study examined characteristics associated with the likelihood of not receiving the COVID-19 vaccine among university staff and faculty during the spring and summer of 2021.

## Methods

### Participants

Participants were staff and faculty at the University of Southern California (USC) in Los Angeles, California. Participants were eligible if they were currently employed at USC, were at least 18 years of age, and provided informed consent.

### Procedure

The USC Institutional Review Board approved the study. Emails were sent to all staff and faculty inviting them to participate in a brief COVID-19 survey and the study was advertised on university websites. Participants provided informed consent electronically and completed the online surveys by clicking on a hyperlink link sent via email. Survey responses were collected between 4/29/2021 and 07/21/2021. After removing duplicate and incomplete surveys, the sample size was 2,125.

### Measures

Demographic variables included self-identified race and ethnicity (White; Asian/Asian American; Black/African American; Hispanic/Latinx; multicultural; or other), sex, age, and political affiliation (Democrat, Independent, Republican, something else). Employment related characteristics included division (staff, faculty, student employee) and subdivision (healthcare related position or all other university division), change in income during the pandemic (decreased or no change/increased) and work-from home (yes or no). COVID-19 history was self-reported and categorized as “yes” or “no”. Additional survey questions focused on housing situations, recent travel outside of Los Angeles, COVID-19 knowledge and attitudes and compliance with prevention behaviors including masking and social distancing. The outcome variable for this study was self-reported vaccination status at the time of the survey.

### Data analysis

Logistic regression models were used to estimate associations and adjust for potential confounders. Variables that were significantly associated with vaccination status at p<0.05 were included in a final multivariable model and those deemed as *a priori* potential confounders (age, sex, and ethnicity). Adjusted odds ratios and 95% confidence interval associated with vaccination status are reported.

## Results

Among the 2817 staff and faculty who were sent invitation emails, 2125 (75.4%) completed the survey and were included in the analysis. Demographic characteristics of the sample are shown in Table 1. Mean age was 42.2 years (±12.3). Most respondents identified as white (40.2%) or Asian / Asian American (23.8%). More than half of participants were female (68.3%) and identified their political affiliation as Democrat (65.6%). Participants were mostly employed as staff (70.5%), worked in a non-healthcare related division (55.7%), had no change in income during the pandemic (72.9%) and currently worked from home (79%). Most participants reported no history of COVID-19 (82.2%) and self-reported that they had received a COVID-19 vaccination [(78.4%) Table 1)].

**Table 1.** Demographic and self-reported characteristics of a sample of faculty and students at a large University in Los Angeles (N=2125)

| Characteristic | N % |
| --- | --- |
| <u>Race/ethnicity</u> |  |
| Asian or Asian American | 501 (23.8) |
| Black or African American | 118 (5.6) |
| Hispanic/Mexican/Spanish/Latinx | 300 (14.2) |
| Multicultural or other | 342 (16.1) |
| White | 846 (40.2) |
| <u>Age group</u> |  |
| 20-32 years | 534 (25.1) |
| 33-40 years | 561 (26.4) |
| 41-51 years | 527 (24.8) |
| 52-85 years | 503 (23.7) |
| <u>Sex</u> |  |
| Female | 1450 (68.2) |
| Male | 674 (31.7) |
| Undefined | 1 (0.01) |
| <u>Division Status</u> |  |
| Staff | 1498 (70.5) |
| Faculty | 469 (22.1) |
| Student employee | 158 (7.4) |
| <u>Subdivision</u> |  |
| Healthcare related division | 941 (44.3) |
| Non-healthcare related division | 1184 (55.7) |
| <u>Change in income</u> |  |
| No change/increased | 1540 (72.5) |
| Decreased | 572 (26.9) |
| Not specified | 13 (0.6) |
| <u>Worked from home</u> |  |
| Yes | 1666 (78.4) |
| No | 443 (20.8) |
| No specified | 16 (0.8) |
| <u>Traveled in/out of Los Angeles</u> |  |
| No | 798 (37.6) |
| Yes | 1327 (62.4) |
| <u>Political Affiliation</u> |  |
| Democrat | 1394 (65.6) |
| Independent | 388 (18.3) |
| Republican | 152 (7.2) |
| Something else | 191 (9) |
| <u>Had COVID-19</u> |  |
| No | 1747 (82.2) |
| Yes | 378 (17.8) |
| <u>Self-reported responses to receiving COVID-19 vaccine</u> |  |
|  | 1665 (78.4) |
| Already received the vaccine |  |
|  | 342 (16.1) |
| Yes, as soon as possible |  |
|  | 34 (1.6) |
| Probably, yes |  |
|  | 20 (0.9) |
| Probably, no |  |
|  | 12 (0.6) |
| No, definitely will not |  |

Adjusted odds ratios and 95% confidence intervals from the multivariable analysis for characteristics associated with the likelihood of being unvaccinated are shown in Table 2. Compared to Non-Hispanic White participants, Asian/Asian Americans (OR=1.44, 95% CI: 1.06, 1.96), Hispanic/Latinx (OR=1.73, 95% CI: 1.21, 2.49) and Multicultural/Other (OR=1.72, 95% CI: 1.24, 2.38) employees had higher odds of being unvaccinated. African American/Black participants were not significantly different than Non-Hispanic Whites. Participants who were older than 32 years had greater odds of being unvaccinated compared to participants in the youngest age quartile (Table 2). Assigned sex and having COVID-19 were not associated with vaccination status. Reporting political affiliation as Independent (OR=1.39, 95% CI:1.04, 1.85) or as something else (OR= 3.84, 95% CI: 2.72, 5.41) were associated with greater odds of being unvaccinated compared to those who self-identified as Democrats. University staff (OR= 1.69, 95% CI: 1.24. 2.30) had higher odds of being unvaccinated when compared to faculty members. Additionally, respondents working in a healthcare related divisions were more likely to be unvaccinated (OR=1.51, 95% CI:1.19, 1.93) compared to employees in non-healthcare divisions. Participants who were unable to work remotely (OR=1.48, 95% CI:1.13, 1.93) and reported a decrease in their income (OR=1.34, 95% CI:1.05, 1.71) also had higher odds of being unvaccinated. Participants who did not travel were more likely to be unvaccinated (OR=1.46, 95% CI: 1.16, 1.83) compared to those who traveled outside of Los Angeles area.

**Table 2.** Characteristics associated with the likelihood of not receiving the COVID-19 vaccine among university staff and faculty using an adjusted multivariate logistic regression model. (N=2125)

| Characteristics | Odds Ratio | 95 % Confidence Interval |
| --- | --- | --- |
| **Race/ethnicity** |  |  |
| White | Reference | Reference |
| Asian or Asian American | 1.44 | 1.06, 1.96 |
| Black or African American | 1.16 | 0.69, 1.95 |
| Hispanic/Latinx | 1.73 | 1.21, 2.49 |
| Multicultural or other | 1.72 | 1.24, 2.38 |
| **<u>Age group</u>** |  |  |
| 20-32 years | Reference | Reference |
| 33-40 years | 2.15 | 1.52, 3.05 |
| 41-51 years | 2.90 | 2.04, 4.12 |
| 52-85 years | 3.44 | 2.37, 4.99 |
| **Assigned sex** |  |  |
| Female | Reference | Reference |
| Male | 1.02 | 0.79, 1.30 |
| **Division Status** |  |  |
| Faculty | Reference | Reference |
| Staff | 1.69 | 1.24, 2.30 |
| Student employee | 1.71 | 0.98, 2.99 |
| **Subdivision** |  |  |
| Non-healthcare related | Reference | Reference |
| Healthcare related | 1.51 | 1.19, 1.93 |
| Income change |  |  |
| No change/increased | Reference | Reference |
| Decreased | 1.34 | 1.05, 1.71 |
| Worked from home |  |  |
| Yes | Reference | Reference |
| No | 1.48 | 1.13, 1.93 |
| Travel in/out of L.A. |  |  |
| Yes | Reference | Reference |
| No | 1.46 | 1.16, 1.83 |
| Political Affiliation |  |  |
| Democrat | Reference | Reference |
| Independent | 1.39 | 1.04, 1.85 |
| Republican | 1.42 | 0.94, 2.31 |
| Something else | 3.84 | 2.72, 5.41 |
| Had COVID-19 |  |  |
| No | Reference | Reference |
| Yes | 1.27 | 0.96, 1.68 |

## Discussion

This study identified characteristics associated with being unvaccinated for COVID-19 among a large, diverse sample of university staff and faculty during the spring and summer of 2021. Survey responses were collected prior to the implementation of a vaccination policy for campus reopening and resumption of in-person learning and activities. We identified several significant characteristics associated with being unvaccinated including race/ethnicity (Asian and Asian American, Hispanic/Latinx and as multicultural), were older than 32 years, unable to work remotely, and political affiliation.

Results from previous studies have shown differences in vaccination rates among races and ethnicities, predominantly among African American and Black participants. [7, 9] Although African American and Black participants in the present study were not more likely to be unvaccinated compared to whites (OR=1.16, 95% CI: 0.69, 1.95), this nonsignificant finding may be due to small sample size (N=118). Our results suggest delayed vaccination is prevalent among many other races and ethnicities especially among participants identifying as Asian/Asian American, Hispanic//Latinx and as Multicultural/Other compared to Non-Hispanic White participants. Participants who have experienced racial discrimination are more likely to be vaccine hesitant, emphasizing the need to address racial disparities as a barrier to COVID-19 vaccination.[16-18]

Reduced vaccine uptake has been associated with socially vulnerable populations, especially among those who suffered a loss in employement and/or income.[19] Financial and employment loss was an unforseeable consequence due to the need to mitigate the spread of COVID-19 resulting in business closures.[20] Reduced or loss of income among populations who were already vulnerable further lowers the likelihood of vaccination.[19] Furthermore, individuals who were unable to take time off of work have cited fear of loss of income or employement as reasons for not being vacinated.[21] Similar results were observed in the present study where prticipants reporting decreased income were more likely to be unvaccinated compared to those who had no change or increase in income. Developing culturally resonant interventions and increased vaccine allocation to more vulnerable populations especially those who face barriers to access because of concerns of reduced income need to be considered as an operational part for future vaccination strategies

The pace at which the COVID-19 vaccine was developed and approved has been associated with delayed vaccination even among healthcare workers.[22, 23] Although Healthcare workers with direct patient contact were priorititzed for early receipt of the COVID-19 vaccine after emergency autorization use by the Food and Drug Assiciation, [24] and despite having potentially greater exposure to COVID-19, many healthcare workers deferred COVID-19 vaccination until more data regarding long-term side effects was available. Are results corroborate this with participants in the present study working in a heathcare associated division of the university had a greater liklihood of being unvaccinated. Although most healthcare workers have greater health related knowledge, it is important to acknowledge and address reasons for vaccine hesitation among healthcare staff.

Political partisanship has been associated with vaccine mistrust and hesitancy among adults in the U.S. [10, 25-27] In the present study, participants identifying as Independent or something else had greater odds of not receiving the COVID-19 vaccine compared to Democrats. Like other barriers to vaccine hesitancy previously discussed in this paper, political beliefs, misinformation and mistrust of science and government needs to be addressed through education.

The current study adds to the literature in several ways. Survey responses were obtained from a large, diverse sample of staff and faculty including healthcare workers. Furthermore, our findings suggest participants who are most likely to be unvaccinated may also have greater risk for exposure such as healthcare workers and those unable to work remotely. Although these results concur with other published literature, to our knowledge this is the first study to investigate behaviors and attitudes towards COVID-19 vaccination among university staff and faculty.

## Limitations

This analysis was based on a non-random sample of university staff and faculty who responded to an online survey and results may not generalize to the larger population or other employees of another university. Future studies are needed to identify correlates of vaccination and vaccine hesitancy among staff and faculty who chose not to participate. Another limitation is self-reported data including COVID-19 vaccination status.

## Conclusions

This study identifies subgroups of university staff and faculty who were unvaccinated for COVID-19 prior to the start of the 2021-2022 academic year. Furthermore, this study provides insight into potential barriers for attaining vaccination and has identified groups that may benefit from targeted education and outreach vaccination campaigns. Additionally, these findings may help generate university policies and programs to address future pandemics.

## Data Availability

Data available upon request

